# Axial Length Matters: Scaling Effects in Retinal Fundus Image Analysis

**DOI:** 10.64898/2026.03.03.26347501

**Authors:** Qingkuo Li, Ajay B Harish, Hongcheng Guo, Jeffrey TW Leung, Hema Radhakrishnan

## Abstract

**Purpose:** Quantitative metrics obtained from retinal fundus images (such as vessel length, tortuosity and other scale-dependent measures) are increasingly used as potential biomarkers for systemic diseases, including cardio- and neurovascular conditions. However, with the increasing prevalence of myopia and related axial growth, this study evaluated whether axial length scaling alters inferred biomarkers compared with measurements obtained without axial length scaling and whether these effects can be corrected.

**Methods:** 2,309 clinic visits from patients aged ≤21 years were analysed for axial-length scaling analysis (range20 to 28 mm). The retinal fundus photographs were automatically segmented using AutoMorph to extract vascular metrics. The parameters were corrected for axial length using correction factors based on the Bennett–Littmann formula and true axial length.

**Results:** Axial length significantly influenced biometric parameters (vessel metrics) derived from fundus photography. The magnitude of error in diameter and length of blood vessels was approximately 4-5% for each 1 mm deviation from the reference axial length of 24 mm, whereas the error in vessel area was approximately 9-10% per 1 mm, consistent with the geometric expectation that area scales with the square of linear dimensions. The scaling corrections for different axial lengths are presented.

**Conclusions:** Axial-length-related magnification introduces systematic bias into retinal vascular metrics from fundus photographs. Bennett–Littmann correction using true axial length reduces these errors and should be adopted in quantitative fundus imaging and AI biomarker development.

**Translational Relevance:** Accounting for axial-length-related magnification may improve the comparability and accuracy of quantitative retinal vascular biomarkers derived from fundus photographs.

## Introduction

Fundus photographs enable non-invasive visualisation of subtle changes in retinal vasculature and neural structures. Artificial intelligence (AI) algorithms based on fundus images are increasingly being used to enable automatic detection of eye diseases such as diabetic retinopathy and automated referral decisions.^1,2^ In addition, deep learning models can predict systemic metabolic indices and cardiovascular risk,^3,4^ as well as systemic diseases such as chronic kidney disease,^5^ with retinal biomarkers identified from fundus photographs.

Retinal biomarkers are also attracting growing interest in the neurological field: structural and vascular alterations in the retina may reflect pathological processes in the brain, and fundus imaging is being explored as an adjunctive tool for the diagnosis of Alzheimer’s disease.^6,7^ In particular, the retinal microvasculature provides a rich set of quantitative features, including vessel calibre, vessel area, vascular density, and branching or bifurcation points. These vascular parameters have been shown to be closely related to multiple disease states and are now being extracted at scale to construct population reference maps of retinal vascular parameters and to elucidate their genetic and phenotypic associations.^8–11^ Collectively, these advances underscore the substantial potential of fundus photography as a “window” onto systemic health.

Geometric measurements derived from fundus images are influenced by ocular magnification, particularly by inter-individual differences in axial length, which directly alter the imaging scale. In simple terms, the longer the axial length (as in highly myopic eyes), the smaller retinal structures appear on the fundus image; conversely, when the axial length is shorter (as in hyperopic eyes), the image is effectively magnified. Classical formulas proposed by Littmann,^12^ Bennett,^13^ and others enable correction for image magnification based on axial length and refractive error, thereby converting image-based measurements into the true physical dimensions of retinal features.

Individualised optical scaling correction is often overlooked in contemporary AI algorithms, and fundus images from different patients are implicitly treated as if they share a common scale. Only approximately 8% of OCTA studies applied an axial length-dependent magnification correction.^14^ Accounting for axial length when analysing parameters such as optic disc morphology and retinal nerve fiber layer thickness in highly myopic eyes show that magnification correction can substantially attenuate or even reverse previously reported thinning trends.^15,16^ Failure to account for axial length– related magnification introduces systematic error, making structures in long eyes appear artificially small and vascular measurements spuriously low.

To our knowledge, no quantitative study has yet established a model describing how this error varies as a function of axial length for vascular biomarkers. The aim of the present study was to apply the Bennett–Littmann magnification correction method to perform axial-length–based scaling of colour fundus photographs from a large cohort of children and young adults (3–21 years). We quantitatively evaluated the magnitude of systematic error in various vascular segmentation metrics that arises when a uniform reference axial length is assumed (i.e. when individualized correction is not performed), and we analysed the functional relationship between this error and axial length to provide quantitative evidence and practical correction strategies for future AI-based analyses of fundus imaging.

## Methods

### 2.1 Study population and data collection

This retrospective study included ophthalmic examination data collected at the Optometry Clinic of The Hong Kong Polytechnic University over a 3-year period (1 January 2022 to 18 July 2025). The study population comprised children and young adults aged ≤ 21 years, as axial elongation typically slows and tends to plateau in adulthood, and this restriction reduces confounding from age-related retinal changes and adult ocular comorbidities. The study adhered to the tenets of the Declaration of Helsinki and was approved by the institutional ethics committee of The Hong Kong Polytechnic University, Hong Kong, China (approval number: HSEARS20250407001). The present study applied a retrospective design and used de-identified clinical records from this previous study.

The final study cohort comprised participants aged ≤ 21 years with at least one eligible imaging event during the study period with both colour fundus photography and an axial length measurement available. Exclusion criteria comprised missing axial length measurements or fundus photographs, invalid or missing date/time fields, as well as extreme axial length values, for example below 18 mm or above 32 mm. The unit of analysis was the imaging event, with each eligible imaging event contributing one data point.

### 2.2 Ocular examinations

All participants underwent comprehensive ocular examinations, including assessment of axial length and refractive status. Axial length was measured using either the IOL Master 500 (Carl Zeiss Meditec AG, Germany) or the AL-Scan (Nidek Co., Ltd., Japan) optical biometer. Previous research has shown excellent agreement between these two instruments.^17^ Refractive error was obtained using non-cycloplegic subjective refraction.

### 2.3 Fundus imaging

Retinal images were acquired using three fundus camera configurations routinely employed at the same centre: Topcon TRC-NW6S (Camera A; 3216 × 2136 pixels) and Topcon TRC-NW8 (Camera B; 3696 × 2448 pixels, and Camera C; 4176 × 2784 pixels; Topcon Corporation, Japan). Camera identity was assigned from the original image resolution. All configurations had a nominal 45° field of view. For each fundus photograph, the horizontal pixel width (denoted W) was extracted and combined with the field of view to compute pixels per degree of visual angle (pixel/deg), which was then used to convert pixel-based measurements into physical units.

### 2.4 Vessel segmentation

Retinal vessel segmentation was performed on the fundus images using the automated retinal vascular analysis software package AutoMorph.^18^ The original colour fundus photographs were input into the AutoMorph pipeline (Figure 1), which applies a series of preprocessing steps and deep learning algorithms to generate a binarised retinal vessel mask and the corresponding vessel skeleton (a one-pixel-wide centreline representation). Although AutoMorph incorporates built-in image quality control and field-of-view cropping, in the present study the pixel width of the original, uncropped images was used to compute the magnification factor (pixel/deg), thereby ensuring that magnification correction was consistently based on the same original image scale.

**Figure 1:**
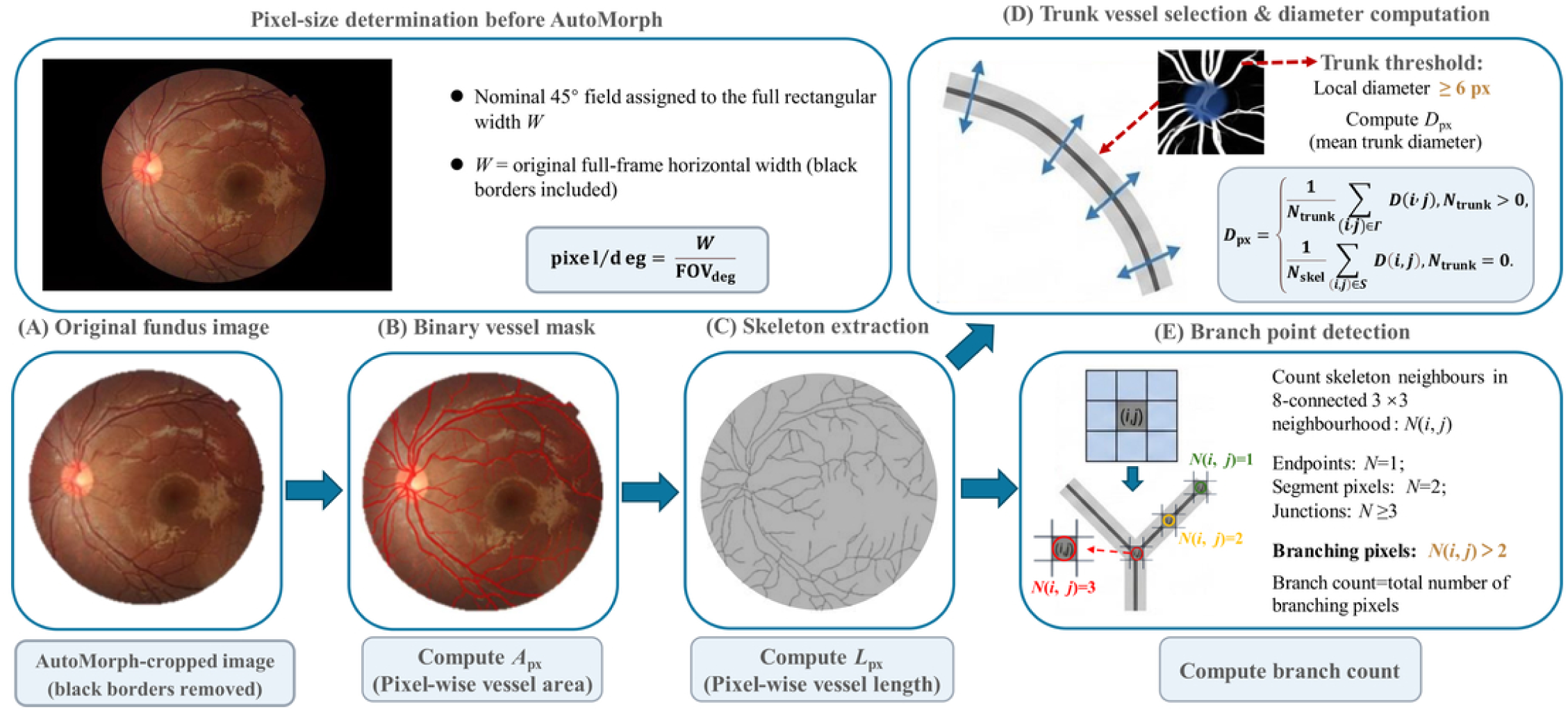
Parameter Definition and Calculation Process. The top panel shows pixel-size determination from the original full-frame fundus image before AutoMorph processing, with W defined as the horizontal image width including the black borders. Panels A–E show (A) AutoMorph cropping with black-border removal, (B) binary vessel mask and vessel-area computation, (C) skeleton extraction and vessel-length computation, (D) trunk-vessel selection and mean diameter computation, and (E) branch-point detection and branch-count computation.

### 2.5 Vascular metrics in pixel space

Binary vessel masks and skeleton maps from AutoMorph were batch-processed using custom Python scripts to derive quantitative vascular metrics in pixel space. The non-zero pixels in the binary mask and skeleton map were treated as vessel and skeleton pixels, respectively.

The vessel area (*A*_px_) was defined as the total number of vessel pixels in the binary mask

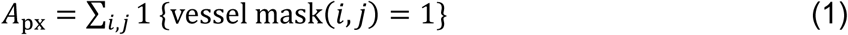

where 1{⋅} denotes the indicator function, which takes the value 1 if the condition in the bracket is satisfied and 0 otherwise.

The vessel skeleton length (*L*_px_) was defined as the total number of skeleton pixels in the skeleton map, representing the cumulative length of the vascular network in pixel units

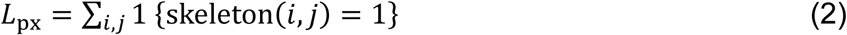

The mean diameter of the major retinal vessels (*D_px_*) quantified the average width of large retinal vessels. A Euclidean distance transform was applied to the binary vessel mask to obtain a distance map *d*(*i*, *j*) giving, for each vessel pixel, its distance to the nearest background pixel. At skeleton locations, this distance was interpreted as the local vessel radius, and the corresponding local diameter was

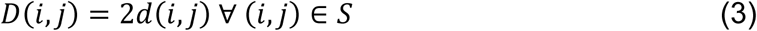

Using descriptive statistics, pixel-wise diameters of major vessel segments were generally >6 pixels. Considering that the set of all skeleton pixels was defined as

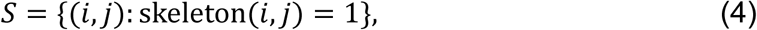

and the trunk-vessel subset be defined using a diameter threshold *τ* = 6 pixels as

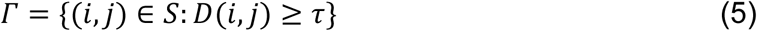

and *N*_trunk_ = |*Γ*|, *N*_skel_ = |*S*| denote the number of pixels in *Γ* and *S*, respectively. Skeleton points with a local diameter ≥ 6 pixels were therefore classified as major vessels, and the mean major-vessel diameter was defined as

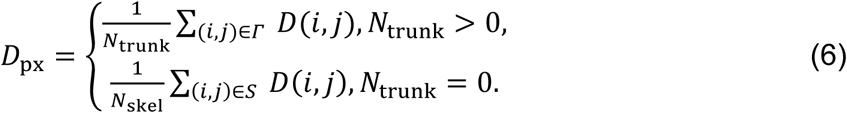

Skeleton points not in *Γ* (i.e. *D*(*i*, *j*) < *τ*) were excluded only from the computation of *D_px_* and were retained for the computation of vessel skeleton length, vessel area and branch count.

Vessel branch count was used to characterise the topological complexity of the vascular network. As shown in Figure 1(E), following skeletonisation, branching points were identified by through the information within the 8-connected neighbourhood in a 3×3 window centred for each skeleton pixel (*i*, *j*). This count was denoted *N*(*i*, *j*). Pixels along an uninterrupted vessel segment typically have *N*(*i*, *j*) = 2, end points have *N*(*i*, *j*) = 1, whereas junction pixels have *N*(*i*, *j*) ≥ 3. Pixels with *N*(*i*, *j*) > 2) were therefore classified as branching points, and branch count was defined as the total number of branching-point pixels.

For each image, the original spatial resolution (width *W* and height *H*) was recorded for use in subsequent calculations of pixel/deg and magnification correction factors.

### 2.6 Axial length–based magnification correction

The number of pixels per degree (pixel/deg) of visual angle, for each fundus image, was defined as

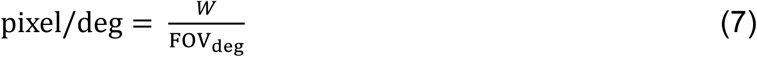

where *W* is the total horizontal pixel width and FOV_deg_ is the nominal 45° field of view. *W* was read directly from the original image file for each photograph, and all magnification corrections were based on the image-specific pixel/deg value.

The influence of axial length on lateral retinal magnification was accounted for using the Littmann–Bennett method to compute the ocular optical magnification factor *q*(*AL*)

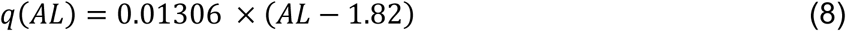

where *AL* denotes axial length (in mm), and the constant 1.82 mm represents the optical distance from the corneal apex to the second principle point of the eye in the Bennett model. This method is widely regarded as the gold standard for estimating ocular magnification.^19^

Considering the true axial length *AL*_true_ of a given eye, the corresponding real-world physical length represented by each pixel was defined as

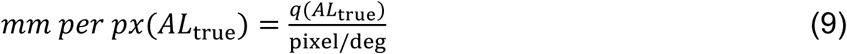

During magnification correction, two spatial scales were defined, namely

#### (1) Reference eye (without-AL-scaling condition), in which a single reference axial length (*AL*_ref_) was assumed for all eyes and the pixel scale was

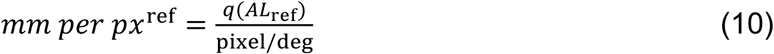

This corresponds to the common practice of using a fixed magnification based on the camera field of view. A reference axial length of 24.0 mm was selected as an approximate reference-eye value based on previous optical eye-modeling work^20^ and was used solely to define the fixed-reference scaling condition.

#### (2) Individualised axial scaling, in which the true measured axial length (*AL*_true_) of each eye was used

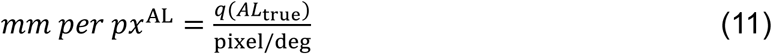

This scale reflects the actual linear size on the retina represented by each pixel after accounting for inter-individual differences in axial length. The vessel diameter, length and area were computed in physical units under both scaling schemes to compare the systematic discrepancies between the with-AL-scaling and without-AL-scaling conditions.

Additionally, the scaling factor (*r*) was defined as

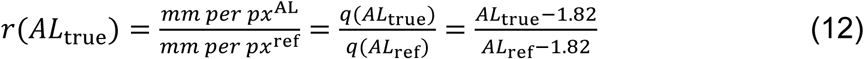

thus, ensuring that the pixel scale factor was determined by the eye’s axial length and independent of the camera’s field of view and resolution.

### 2.7 Conversion of metrics to physical units

The *mm per px*^ref^, and *mm per px*^AL^ were employed to convert all vascular metrics, defined in pixel space, into physical quantities expressed in millimetres or square millimetres. *D*_px_, *L*_px_, and *A*_px_ denote, respectively, the mean trunk-vessel diameter, total vessel skeleton length, and vessel area of a given fundus photograph in pixel space.

Considering the no-scaling (reference axial length) condition, the corresponding physical quantities were computed as:

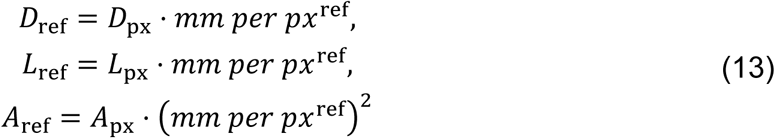

and under the individualized axial-length scaling condition, the corresponding physical quantities were:

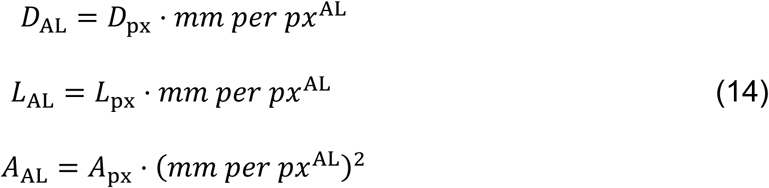

Vessel branch count reflects only the topological structure of the vascular network and is independent of geometric magnification; its value therefore remains unchanged across scaling conditions.

### 2.8 Error definition and statistical analysis

The systematic biases introduced by ignoring axial length-related magnification were quantified by considering the measurements obtained under individualised axial-length scaling as the reference ground truth, and metrics derived under the uniform reference axial-length assumption as uncorrected values. Percentage errors for trunk-vessel diameter, vessel skeleton length and vessel area were defined as:

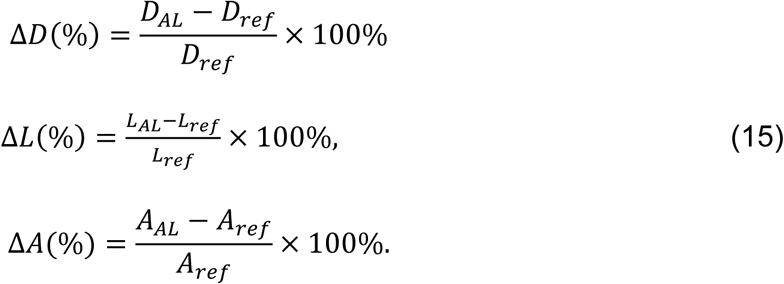

The analytical expressions for these percentage errors were derived using scaling factor (*r*) as

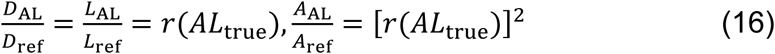

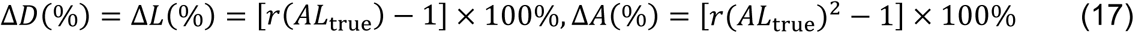

Regression models were used to investigate the quantitative relationship between magnification error and axial length. Considering Δ*D*(%) as the dependent variable and axial length (*AL*_true_) as the independent variable, a linear model

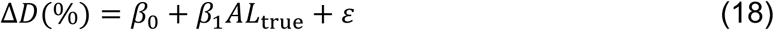

with parameters (*β*_0_, *β*_1_, *ε*) are considered for evaluation. Similarly, regression models were fitted for Δ*L*(%) and Δ*A*(%). Given the expected quadratic dependence of Δ*A*(%) on *AL*_true_, a second-order polynomial term was additionally included when modelling Δ*A*(%). The primary focus of this study was on testing the variation of the slope parameter *β*_1_ w.r.t zero as an estimate to determine whether magnification error exhibited a systematic trend with axial length.

## Results

### 3.1 Study population

The original dataset had 2,666 visit records, with a mean age of 10.56 ± 4.75 years, a median age of 9.72 years, and a range of 2.89∼44.05years. Following the filtering procedure, the filtered dataset comprised 2,394 visits, with a mean age of 10.99 ± 4.71 years, a median of 10.08 years, and an age range of 3.17-44.05 years. This filtered dataset was further reduced to only include patients of ages≤21 years. In the final dataset (≤21 years) used for magnification-correction analyses, the 2,309 visits had a mean age of 10.47 ± 3.35 years, a median of 9.95 years, and an age range of 3.17-20.67 years. Full cohort characteristics and variable-specific data availability, including SER measurements, are provided in Supplementary Table 3.

### 3.2 Impact of axial length on scaling factor

Following the procedures described in Section 2.6, the pixel-space metrics for vessel diameter (*D*_px_), length (*L*_px_) and area (*A*_px_) are obtained. For each image, the axial-length–based physical scale mm per pixel^AL^ was computed deterministically using the image-specific pixel/deg and the measured *AL*_true_ as defined in Section 2.6.

As shown in Figure 2, the 4,631 eligible images were stratified according to their original image resolution: Camera A, 3216 × 2136 pixels (n = 1,076); Camera B, 3696 × 2448 pixels (n = 2,387); and Camera C, 4176 × 2784 pixels (n = 1,168). For each camera group, the scatterplots of mm per pixel^AL^ were plotted against the true axial length *AL*_true_ and followed the expected linear relationship. Across all three original image-width groups, with-AL-scaling physical pixel size increased monotonically with axial length, and given as

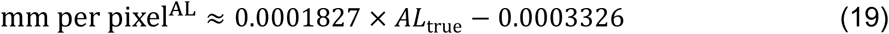

**Figure 2:**
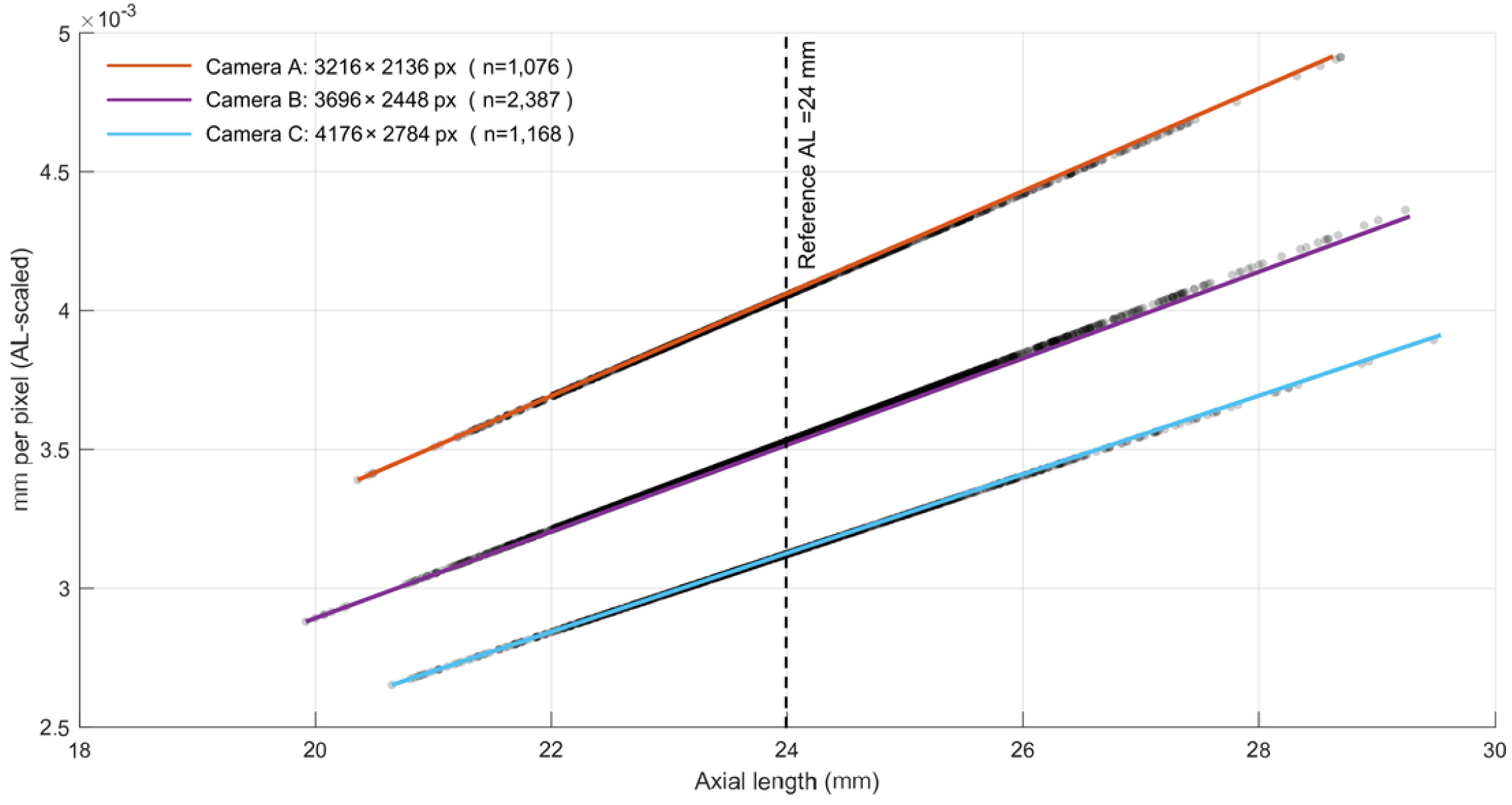
Relationship between axial length and physical pixel size (mm/pixel) stratified by the horizontal width (W) of the three fundus cameras used: Camera A (W = 3216 pixel, top line), Camera B (W = 3696 pixel, middle line), and Camera C (W = 4176 pixel, bottom line)

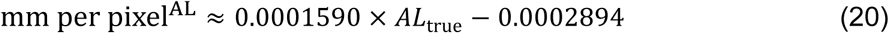

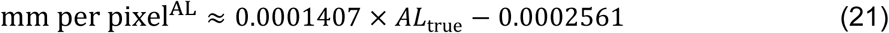

### 3.3 Effect on vessel metrics

The bias introduced by omitting axial-length correction, defined in Section 2.8, is qualitatively assessed using the relative errors between the reference-eye and individualized-scaling conditions (*ΔD*(%), *ΔL*(%), *ΔA*(%)).

Considering the Bennett formula and fixing *AL*_ref_ = 24.0 mm, the magnification factor can be written explicitly as:

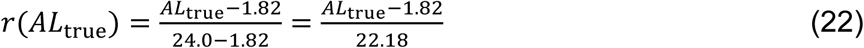

and the relative errors as

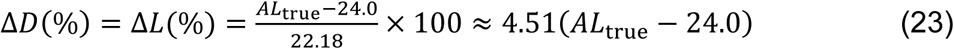

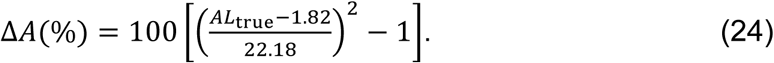

It follows from these expressions that when *AL*_true_ < 24 mm, Δ*D*(%) and Δ*L*(%) are negative, indicating that using 24 mm as a universal reference leads to systematic overestimation of vessel diameter and length in eyes with shorter axial length.

Conversely, when *AL*_true_ > 24 mm, vessel dimensions are systematically underestimated, and the area error Δ*A*(%) increases approximately quadratically with axial length.

Figure 3 illustrates the dependency between axial length and trunk-vessel diameter, total vessel skeleton length, and vessel area, and shows representative comparisons without-AL-scaling values (*AL*_ref_ = 24.00 mm; *D*_ref_*, L*_ref_*, A*_ref_) and with-AL-scaling values (using measured *AL*_true_; *D*_AL_, *L*_AL_, *A*_AL_). In eyes with shorter *AL*_true_, the without-AL-scaling measurements of diameter, length and area were systematically higher than the with-AL-scaling values; whereas in eyes with longer *AL*_true_, the opposite pattern was observed. Around the reference axial length (*AL* ≈ 24 mm), the discrepancy between the two methods is minimal, as expected.

**Figure 3:**
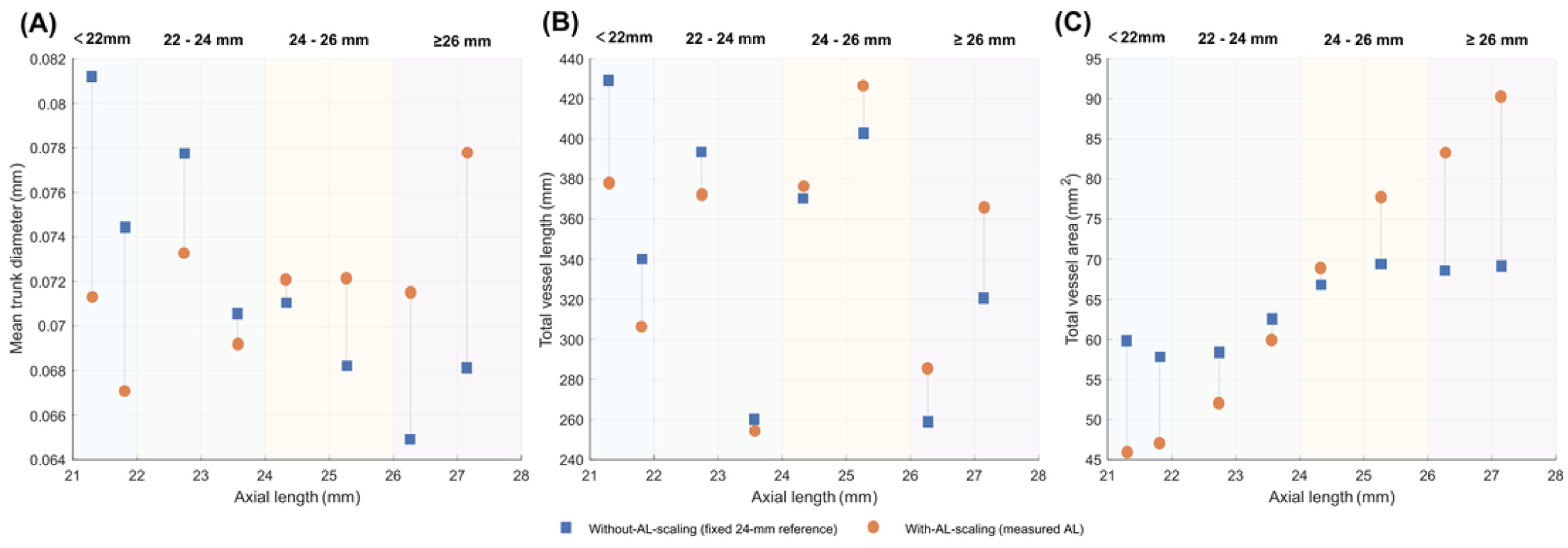
Comparison of retinal vascular metrics with and without AL scaling in representative eyes. Axial-length strata were defined as AL<22 mm, 22≤AL, 24 mm, 24≤AL, 26 mm, and AL≥26 mm. Two representative eyes were selected from each stratum. Panels show (A) mean trunk-vessel diameter, (B) total vessel skeleton length, and (C) total vessel area. Blue squares indicate values obtained without individualized axial-length scaling using a fixed 24-mm reference, and orange circles indicate values obtained with scaling based on the measured axial length. Grey connectors link paired values from the same eye.

In addition, linear regression was used to determine the relation between the axial length (*AL*_true_) and Δ*D*(%)and Δ*L*(%) against using all eligible observations, as shown in Figure 4. The fitted models yielded identical slopes of 4.51% per mm with an intercept of approximately −108.2, consistent with the analytic expressions above. Accordingly,

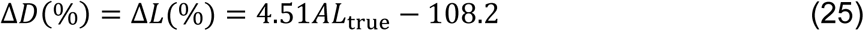

**Figure 4:**
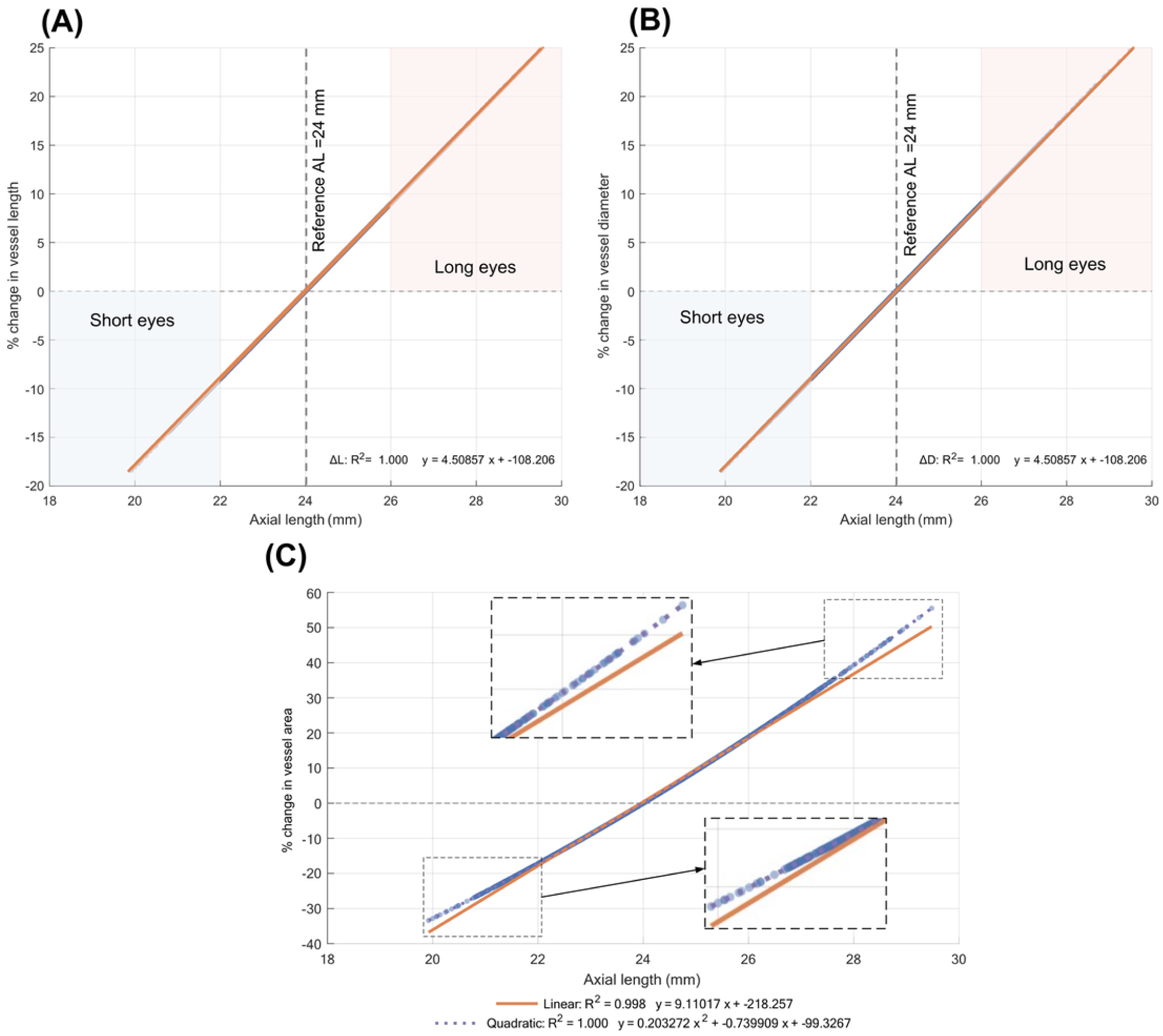
Axial-length dependence of magnification-related error in retinal vascular metrics when axial-length scaling is omitted. (a) ΔL (%) vs. AL (b) ΔD (%) vs. AL (c) ΔA(%) vs. AL (Degree1 & Degree-2); ΔD(%), ΔL(%), and ΔA(%) are plotted against *AL*_true_, where Δ denotes the percentage difference between measurements computed without-AL-scaling using a fixed reference *AL_ref_*=24.0 mm and those computed with *AL*_true_-based scaling. Solid lines indicate linear regression fits in (a–b); in (c), both linear and second-order polynomial fits are shown, with model equations and *R*^2^ reported on the plots

In accordance with the standard assumption that area-based error scales with the square of linear magnification, Δ*A*(%) was modelled using a second-order polynomial in *AL*_true_. The quadratic model provided an excellent fit (R² =1.00) and yielded:

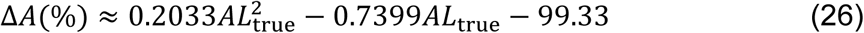

The imaging events were classified into four axial-length groups: *AL*_true_ < 22 mm, 22 ≤ *AL*_true_ < 24 mm, 24 ≤ *AL*_true_ < 26 mm, and *AL*_true_ ≥ 26 mm to enable and support analysis. For each group, the mean and standard deviation of *D*_ref_, *D*_AL_, and Δ*D* (%) were calculated, as well as the corresponding metrics for vessel length and area (*L* and *A*) (see Supplementary Table 1). Overall, for *AL*_true_ < 22 mm, vessel diameter and length were overestimated by approximately 8-10% in the uncorrected condition, and vessel area was overestimated by about 17-20%. In contrast, for *AL*_true_ ≥ 26 mm, diameter and length were underestimated by roughly 10-12%, and area by about 22-25%.

As illustrated in Figure 4, across the full range of axial length, the magnitude of error in diameter and length was of the order of 4–5% for each 1 mm deviation from the reference axial length *AL*_ref_ = 24 mm, whereas the error in area was approximately 9– 10% per 1 mm deviation, consistent with the geometric expectation that area scales with the square of linear dimensions.

Another pertinent metric often used in disease classification is the branch count. Computing the branch count in the dataset, using the method illustrated in Figure 1, the branch count values in the original segmentation output and in the magnification-corrected results were identical for every image. The linear regression of branch count on *AL*_true_ yielded *R*² ≈ 0.003, indicating virtually no explainable linear relationship between the two.

## Discussion

Recent vascular biomarker studies, based on fundus images, using AI as yet do not incorporate individual axial-length information into their analytical pipelines. The results of this study demonstrate that axial length correction of retinal images is needed to counteract systematic, axial-length-dependent errors into vessel diameter, length and area. Linear metrics showed approximately proportional changes with axial length, whereas area-based metrics showed a higher-order dependence; in contrast, branch count, as a topological measure, showed no material association with axial length.

Most prior work on magnification correction with axial length has focused on optic disc morphology, retinal nerve fibre layer (RNFL) thickness, and OCT/OCTA-derived structural parameters. Early studies^19, 13^ showed that, without correcting for axial length–related magnification, optic disc area can be systematically under- or overestimated. More recently, Akiyama et al.^15^ demonstrated that, after applying axial length-based magnification correction, the apparent thinning of peripapillary capillary density and RNFL thickness with increasing axial length is partially or completely abolished, suggesting that some previously reported “myopia-related structural changes” may largely reflect image-scaling artefacts rather than true tissue alterations. At the vascular level, Yii et al.^16^ reported that each 1 mm increase in axial length was associated with a reduction of 0.5–0.9 pixels in retinal arteriolar and venular calibres; once Bennett–Littmann correction was applied, this negative association essentially disappeared.

Compared with these studies, the present work differs and extends the literature in two important ways. First, the emphasis on segmentation-based vascular metrics expands the correction to network-level indices derived from automated segmentation, including total vessel area, skeleton length, and mean trunk-vessel diameter. Such metrics are widely used as AI biomarkers and in population-based epidemiological studies and for automated disease classification, yet there has been little empirical work in systematically quantifying their sensitivity to axial length–related magnification. As vascular calibre and network morphology are used in automated screening and disease classification for conditions such as hypertensive and diabetic retinopathy, correcting vascular metrics for axial length is likely to reduce eye-size–related magnification bias and improve interpretability. Secondly, this work provides an explicit functional description of the error as a function of axial length; linking ΔD(%), ΔL(%), and ΔA(%) to *AL*_true_, and validating these relationships using stratified summary statistics. This relationships can be directly incorporated into engineering and analytical pipelines to quantify expected bias and to report scenario-based error when individual axial length can only be estimated and measurements are unavailable. Functional relationships provided in this work, linking magnification error to axial length, together with the stratified error summaries, can be used to construct simple “error look-up tables” or “correction-factor modules.” When individual axial-length measurements are unavailable, but the axial-length distribution of the cohort is known, allowing estimation of the order of magnitude of potential bias. When individual axial-length data are available, a Bennett–Littmann correction module can be embedded directly into the AI pipeline to achieve normalization of physical scale.

Retinal vascular morphology is used, in risk stratification for systemic diseases. Narrower retinal arterioles and wider retinal venules have been linked to coronary heart disease and other adverse cardiovascular outcomes, and retinal microvascular signs are increasingly seen as markers of overall vascular health.^21^ In large population cohorts, retinal vessel calibre measured automatically using deep learning has also been linked to a higher risk of myocardial infarction and can add useful information to traditional risk models.^22^ In addition, AI models based on fundus photographs have been used to predict major adverse cardiovascular events, suggesting that an integrated approach using retinal images for cardiovascular risk assessment is feasible.^23,24^ In neurovascular and neurodegenerative research, population-based studies have also linked retinal vessel calibre to dementia risk, and venular widening and small-artery changes have been associated with vascular dementia and related outcomes.^25–27^ These findings support the use of automated vessel calibre assessment and deep learning on fundus images in cardiovascular and brain health prediction pipelines.^28^

The present findings indicate that, if axial-length-related magnification is ignored, scale-dependent vascular measures, including mean vessel diameter, total vessel skeleton length, and vessel area, will be systematically underestimated in eyes with longer axial lengths and overestimated in eyes with shorter axial lengths. In risk prediction models that treat reduced vessel diameter or reduced vascular density as a higher risk pattern, this geometric bias may lead to systematic calibration errors across refractive groups.

For example, if a model uses arteriolar narrowing as a sign of higher cardiovascular risk, myopic eyes with longer axial lengths may appear to have narrower or sparser vessels when magnification is not corrected, which may increase the chance of false-positive risk attribution. Conversely, hyperopic eyes with shorter axial lengths may show bias in the opposite direction, potentially masking the risk. This interpretation is in line with large cohort evidence showing that arteriolar narrowing is associated with a higher risk of myocardial infarction,^22^ and with systematic review findings linking retinal microvascular changes to coronary heart disease risk.^29^ Some studies have also suggested that retinal vascular density and related measures are associated with coronary atherosclerosis burden, further supporting the use of vessel thinning or reduced vessel density as risk signals.^30^ This potential confounding is especially important in multi-centre or population-based screening, because axial length distributions can differ across subgroups. For these reasons, when developing and validating fundus-based biomarkers and AI risk models, Bennett-Littmann correction should be used whenever axial length data are available, and stratified calibration or sensitivity analyses should be considered when such data are missing. Results should also clearly state that axial length and magnification can cause systematic shifts in scale-dependent vascular measures and their risk interpretation, to reduce the risk of misclassification in myopic and hyperopic populations.

## Data Availability

All data directly produced in the current study are available upon reasonable request to the authors.

## Author Contribution Statement

**QL** was responsible for Software development, Investigation, Formal analysis, Writing – original draft; **ABH for** Conceptualization, Funding acquisition, Project administration, Methodology, Resources, Supervision, Writing – review & editing; **HG and JL** for Data curation, Clinical image data acquisition; **HR** for Conceptualization, Resources, Supervision, Writing - review & editing.

